# Factors Influencing the Implementing Readiness of Shared Medical Appointments in China’s Primary Healthcare Institutions: A Mixed-Method Study Utilizing Qualitative Comparative Analysis

**DOI:** 10.1101/2024.06.19.24309131

**Authors:** Wei Yang, Lingrui Liu, Jiajia Chen, Run Mao, Tao Yang, Lang Linghu, Lieyu Huang, Dong (Roman) Xu, Yiyuan Cai

## Abstract

**Background and Objective** Diabetes mellitus (DM) is a mounting public health concern in China, home to the largest number of patients with diabetes globally. A primary challenge has been the integration of high-quality chronic disease services, with poor outcomes and inefficient health management intensifying the disease burden. Shared Medical Appointments (SMAs) offer a promising solution, yet evidence of their practical application in resource-limited settings like China’s primary healthcare institutions is scant. This study aims to evaluate the organizational readiness for change (ORC) in implementing SMA services in Guizhou province’s primary healthcare institutions and to identify determinants of high-level ORC to foster implementation success.

**Methods** This study employed a mixed-method approach. The validated Chinese version of the Workplace Readiness Questionnaire (WRQ-CN) was used to assess the ORC status across 12 institutions participating in the SMART pilot trial. A Normalization Process Theory (NPT) -guided qualitative interview and quantitative survey were used to collect the conditions. Data analysis encompassed standardized descriptive statistics, Spearman correlation analysis, and qualitative comparative analysis (QCA) to discern condition variables and configurations that are favorable to high-level ORC.

**Results** The study engaged 70 institutional participants, including administrators, clinicians, and public health workers. The median ORC score was 105.20 (101.23-107.33). We identified 12 condition variables through the interview and survey. The Spearman correlation analysis highlighted a moderate correlation between Specific tasks and responsibilities (r=0.393, p=0.206) and Key participants (r=0.316, p=0.317) with ORC. QCA also revealed these condition configurations and pathways that collectively align with heightened ORC, accentuating the pivotal role of key participants.

**Conclusions** This study unveiled a spectrum of dynamic conditions and pathways affecting ORC, which are consistent with the NPT-based theoretical steps. They were essential for attaining high-level ORC in rolling out health service innovations like the SMART study, especially in resource-limited settings.

---

Healthcare services innovation; Mixed-methods study; Qualitative comparative analysis (QCA)

- **What is already known on this topic** - Organizational Readiness for Change (ORC) has been extensively evaluated within implementation research in developed countries. However, there was a significant knowledge gap in understanding and promoting ORC in low and middle-income countries (LMICs) and resource-limited communities. Current studies primarily employ either qualitative interviews or quantitative methods to analyze ORC, neglectiong the the advantages of mixed methods in comprehensively exploring ORC and its influencing factors.
- **What this study adds** - This study pioneers the exploration of ORC within primary healthcare settings in low-resource settings through a QCA-based mixed method. It fills a critical gap in the literature by providing insights into fostering ORC in contexts previously underexplored.
- **How this study might affect research, practice or policy** - The study illuminates specific pathways to improve ORC in primary healthcare contexts. The mixed method significantly enhances the depth and breadth of our understanding, offering an NPT-based dynamic perspective on the factors contributing to the readiness to implement health service innovations.

## Background

Diabetes mellitus (DM) represents a significant public health challenge <u>globally,</u> and serving as a major contributor to morbidity and mortality rates, with its prevalence rapidly increasing and imposing a considerable disease burden worldwide^1^. The prevalence of type 2 diabetes Mellitus (T2DM) has surged within Chinese populations^2^. Specifically, China has the largest number of patients with diabetes (PWD), accounting for 1 in 4 of all adults living with diabetes worldwide. (140.9 million)^3^. The lack of integrated and high-quality chronic disease services has led to poor patient outcomes and inefficient health management^4^, thereby exacerbating the overall disease burden^5^. In response, China launched the "National Essential Public Health Services Program" (*NEPHS* Program) in 2019, aimed at offering essential, complementary diabetes management services in communities^6^. However, many resource-limited communities encountered numerous challenges, such as inadequate coordination between public health and curative services, lack of collaboration among healthcare implementation practitioners, insufficient physician-patient communication, low healthcare quality, and a deficiency in service^6^ ^7^.

The Shared Medical Appointment (SMA) service, a patient high-level engagement approach, delivers diagnosis, treatment, and health management services simultaneously to a group of pre-scheduled appointments with patients with similar clinical conditions^8^ ^9^ in outpatient care settings by an interdisciplinary team of healthcare professionals. The SMA services were developed based on the Chronic Care Model (CCM)^10^, which has been demonstrated for its positive effect on integrated treatment and health management, enhanced patient health outcomes, and increased service efficiency in developed countries and well-resourced settings^11^ ^12^. However, evidence of practical experience in resource-limited communities remains scarce^13^. To address this gap, the Shared Medical Appointment for patients with diabetes in communities in Guizhou province in China (i.e., *SMART study*) was undertaken to select the optimal set of SMA service components, including one-on-one or group-based consultation and online or offline health education services, an innovative service model in resource-constrained community health centers (CHCs) and township health centers (THCs) in China^14^. As part of this trial program, this study focuses on evaluating the institution’s organizational readiness of change (ORC) before initiating the *SMART study* pilot trial and identifying variables for high-level ORC to enhance the implementation process.

An accurate assessment of ORC could provide information about staff’s commitment and efficacy to innovation (e.g., changing of service model), which in turn predicts the likelihood of successful innovation implementation^15^. When an institution’s ORC was good, employees would invest more effort into innovation and be willing to overcome obstacles and setbacks^16^. An inadequate organizational readiness to implement innovation will hinder the implementation following the protocol, leading to an overdue or even failed implementation^17^. Despite ORC’s importance, readiness before implementing innovation and influencing factors in resource-limited primary care institutions have been under-researched^18^.

Current explorations of ORC have focused on developed countries^19^, leaving a knowledge gap in understanding how to foster ORC in low- and middle-income countries and resource-limited communities settings. Meanwhile, previous studies have typically analyzed ORC solely using qualitative interviews or quantitative surveys, and overlooking the benefits of mixed-methods in comprehensively exploring the configuration of ORC and its influencing condition variables^19^.

Organizational innovation in primary care practices faces a lot of challenges and complexities^20^. The Normalization Process Theory (NPT) offers a theory and framework for organizing these challenges into four consecutive phases relevant to innovation implementation^21^. We hyphosis that achieving the high-level ORC status was dynamic and interconnected (Appendix 1-A). The readiness for implementation would shaped by practitioners’ initial understanding of the *SMART study*’s relevance and significance (i.e., *Coherence*) when it was first introduced. Subsequently, their engagement and mental investment in the program (i.e., *Cognitive participation*) played a crucial role in achieving a high-level readiness for the innovation prior to its actual implementation^22^ ^23^. This study employs the NPT to develop an interview outline to identify challenges that achieve high-level ORC. Furthermore, we used Qualitative Comparative Analysis (QCA) to conduct a rigorous exploration of the essential factors for the implementation of the *SMART study* in primary care settings in Guizhou, China. The QCA approach leverages both qualitative and quantitative data to analyze the interplay among various concurrent influencing factors (i.e., condition variables) across multiple cases (i.e., institutions). These conditions, whether individually or in combination, may correlate with a high-level ORC (i.e., outcome variable).

Thus, we aimed to combine qualitative and quantitative data and elucidate the multifaceted relationships between various condition variables and their collective impact on achieving high-level ORC, thereby facilitating the successful rollout of the *SMART study*. It would provide a reference for subsequent improvement in preparing for innovation implementation.

## Methods

This study utilized a mixed-methods approach^24^ to find and analyze the condition variables and pathways that would be most conducive to high-level Organizational Readiness for Change (ORC). This study’s dual-faceted methodological design enabled a comprehensive understanding of the context of high-level readiness, thereby contributing to the evidence base on effective implementation strategies within organizational settings.

### Conceptual Frameworks

We developed an interview outline based on the NPT^25^ ^26^ to explore the likely ORC-associated condition variables. To assess the ORC status of the organization by utilizing the localized and validated Chinese version of the Workplace Readiness Questionnaire (WRQ-CN), which was developed according to Weiner’s Organizational Readiness for Change theory ("ORC Theory"). Detailed information on the validation of WRQ-CN will be published separately. Appendix 1 depicts the NPT-informed interview outline.

### NPT operationalization

As shown in Appendix 1-A, the NPT encompasses four interactive constructs^27^ ^28^. We regarded the four constructs as the preparation phase and implementation phase separately, each phase including two steps. We operated them as follows:

The innovation begins with participant *Coherence*, i.e., the organization’s individual and collective psychological readiness before innovation begins. *Cognitive Participation* follows the implementation practitioners’ preparatory stage for implementing innovation; once readiness is adequate, *Collective Action* is taken to operationalize the innovation in the organization. Finally, *Reflexive monitoring occurs, and* based on the implementation experience and outcomes, the organization assesses the impact of the implementation of the innovation on itself. It makes corresponding adjustments to better adapt to the innovation. The interview outline was presented in Appendix 2-B.

The study utilized the first two constructs of NPT, *Coherence* and *Cognitive Participation*, prior to initiating the *Collective Action* phase to inform the development of an interview guide to map the likely condition variables that may associated with ORC in implementing public health interventions. The *Collective Action* and *Reflective Monitoring* constructs were not employed at this stage because they are more applicable in the post-implementation of the *SMART study*. Thematic Analysis (TA)^29^ was used to analyze the interview texts to identify the condition variables impacting ORC ahead of the *SMART* pilot trial. Following a standardized analysis approach, a set of variables was constructed. Based on the set of variables derived, we developed a quantitive-oriented questionnaire of influencing condition variables (i.e., Influencing Factor Quantitative questionnaire, IFQ questionnaire). Each survey question item was rated on a six-point scale: 0 = indicating no influence, 1 = very low, 2 = relatively low, 3 =moderate, 4 = high, and 5 = very high influence. Scores on the higher end of the scale indicated a greater influence of the variable.

### ORC Theory operationalization

Peggy et al.^30^ ^31^ developed the Workplace Readiness Questionnaire (WRQ) drawing from the ORC Theory. The WRQ consists of five dimensions and 32 items: *change context, change valence, information assessment, change commitment, and change efficacy*. The definition of each domain and its composed items was presented in Appendix 2.

We translated and contexted into a Chinese version, with the content validation conducted to validate the scale (The scale items achieved conceptual, linguistic, and semantic equivalence between English and Chinese. Two rounds of Delphi studies revealed that the item-content validity index (I-CVI) ranged from 0.73 to 1.00, kappa values were between 0.70 and 1.00, and Kendall’s W coefficient was 0.881. The scale-content validity index (S-CVI) was *≥*0.97, indicating good validity of the scale items and overall content, with a high degree of consensus among experts. The results of face validity verification showed that the importance of each item was more significant than 1.5, indicating that the scale *’* s appearance, content, or format can effectively measure the intended content. The detailed methods and results of the Chinese version of WRQ will be published separately). The Chinese version WRQ uses both the Likert-5 scales (1=Never/Strongly Disagree, 2=Rarely/Disagree, 3=Sometimes/Neutral, 4=Often/Agree, 5=Always/Strongly Agree) and binary scores (1=Yes, 0=No).

### Settings

This study was conducted in the same settings as the *SMART* pilot trial^14^. The *SMART study* purposely selected Bozhou and Bijiang districts as the research settings, where GDP per capita was comparable to that of countries such as Bosnia and Herzegovina(US$7,585) and Dominica(US$ 8,414) in 2022^32–34^. These include community health centers in the urban areas and township health centers in the rural areas, encompassing both publicly and privately owned entities. These centers were integrated within the local government’s diabetes management program, typically through government service contracts. Bozhou and Bijiang comprise 40 primary care or township health centers. In the pilot phase of the *SMART study*, we purposely selected 12 centers, eight of which were from Bozhou and four from Bijiang. Six were from resource-limited settings, and others were from relatively better-resourced settings in two areas (Appendix 3).

### Participants

The study included three types of participants. They were the institution’s administrators, clinical doctors responsible for patients with T2DM treatment, and public health doctors responsible for the patient’s health management. These participants were engaged through both qualitative interviews and quantitative surveys, with at least one participant from each participating primary healthcare facility.

### Data collection

The data collection process was separated into two continuous phases:

**Phase 1:** participant qualitative interviews. First, the NPT-based interview outline will be utilized to conduct the group interviews with three types of participants. The interviews were conducted at the *SMART* pilot trial launching and training sessions, which spanned approximately 60-90 minutes for each participant category. At the end of the interview, the participants were asked how many days they would need to prepare for the conduction. A common preparation period cited by the participants was 14 days. During this interval that the institutions were preparing for the launching, the researchers transcribed the interview text, conducted coding and analyses following the standard TA approach using the Nvivo software (version 12.0)^35^, which informed the development of the Influencing Factor Quantitative questionnaire (IFQ questionnaire).

**Phase 2:** Quantitative assessment of the condition variables condition variables impact and readiness status. All participants of the *SMART* pilot trial were surveyed using a quantitative investigation (i.e., a "Full-Sample" survey), including the WRQ-CN, IFQ questionnaires, and participants’ characteristic information.

Data were collected and administered using the Research Electronic Data Capture platform (REDCap)^36^.

### Data analysis

This study used thematic analysis (TA) to summarize the interview texts and statistically characterize the derived condition variables. *Spearman* correlation analysis was conducted to assess the degree to which these condition variables were correlated. Additionally, recognizing the complexities in practices and the nature of the primary healthcare settings, where multiple pathways (i.e., a combination of multiple conditions) could lead to the optimal ORC, we employed Qualitative Comparative Analysis (QCA) to examine the cross-case patterns for key condition variables that would be most conducive to high-level ORC, facilitating the effective implementation of the *SMART* intervention. We used fuzzy-sets QCA (fsQCA) to address hierarchical variables without dichotomizing or multi-classifying the variables, which can preserve the information and continuity of the condition and outcome variables in this study^37^ ^38^.

**1) Statistical characterization and *Spearman* correlation analysis.** We started with a standardized descriptive analysis and *Spearman* correlation analysis to characterize the condition variables and their correlations. When the correlation value (*r*) was equal to or more than 0.3, it presented a moderate level of correlation^39^, with the scatter diagram plots generated for visual interpretation.
**2) Variables calibration and membership calculation.** Utilizing the direct calibration, we transformed the raw scores into a rescaled format ranging from 0 to 1 for both condition variables and outcome variables using the 90th, 50th, and 10th percentiles, measuring the extent to which this condition (i.e., factor, or, variable) was met for each primary healthcare facility site, which was the unit of analysis. A score of 0 indicates the condition was not fully membership, while a score of 1 indicates the condition was fully membership. For any variables with a rescaled score of 0.5, we incrementally adjusted this score to 0.501^40^, thus conferring that this condition was fully met for the site.
**3) Necessity and coverage analysis.** We first identified the necessary conditions for "high-level ORC" in the primary healthcare facility sites. Then we examined the sufficient conditions while meeting the goodness-of-fit criteria (i.e., a consistency score higher than 0.9, indicating a strong association between the conditions and outcomes^41^).
**4) Truth table construction and configuration analyses.** We constructed a truth table and performed configuration analyses to identify the cross-case patterns and the combinations of conditions that could be most conducive to a high-level ORC. Following Ragin et al.’s guidelines for selecting the number of conditions in QCA^41^, we chose four to seven condition variables when the case number ranges from 10 to 40. The solution consistency score of 0.8 or more was considered to be a sufficient condition for achieving the ORC result.
**5) Robustness test.** We conducted robustness tests using two methods. 1) We changed the consistency threshold from 0.85 or 0.90. 2) We added other variables related to the results for the robustness test to ensure the rationality and accuracy of the results^42^ ^43^.
**6) Interpret result.** Visualize and interpret the results of the pathways (e.g., the combinations of conditions) . A pathway flow between influence factors and high-level ORC was drawn after we further analyzed by combining frequently emerging conditions in different NPT domains.

This study followed the Mixed Methods Article Reporting Standards (MMARS) and the Strengthening Reporting of Observational Studies in Epidemiology (STROBE)^44^ ^45^.

A detailed description of the operational use of QCA in this study was available in Appendix 4 and Appendix 5.

## Results

### Characteristics of participants

In the study, 70 participants were interviewed, comprising 38.57% males (n=27) and 61.43% females (n=43). The majority (68.57%) were under 40 years of age, with a predominant education level of a bachelor’s degree (58.57%). Most participants had less than ten years of work experience (64.29%). Professionally, 37.14% were physician assistants or technicians, and 30% were attending physicians, primarily in clinical medicine (34.29%) and nursing (17.14%) specialties (Table 1).

**Table 1.** Characteristics of participants

| Categories | n | Proportion (%) |
| --- | --- | --- |
| Gender |  |  |
| Male | 27 | 38.57 |
| Female | 43 | 61.43 |
| Age (years) |  |  |
| ≤30 | 27 | 38.57 |
| 30~ | 21 | 30.00 |
| >40 | 22 | 31.43 |
| Educational background |  |  |
| Secondary vocational school education | 3 | 4.29 |
| Three-year college Education | 26 | 38.57 |
| Bachelor degree | 41 | 57.14 |
| Work experience (years) |  |  |
| ≤10 | 45 | 62.86 |
| 10~ | 8 | 10.00 |
| >20 | 17 | 24.29 |
| Job title |  |  |
| No | 16 | 22.86 |
| Physician assistants/technicians (junior) | 26 | 37.14 |
| Attending physician (intermediate) | 21 | 30.00 |
| Associate Consultant/Deputy Chief Physician (senior) | 7 | 10.00 |
| Specialty |  |  |
| Clinical medicine <sup>a</sup> | 26 | 37.14 |
| Nursing | 11 | 15.71 |
| Traditional Chinese medicine | 9 | 12.86 |
| General Practice | 9 | 12.86 |
| Integrative Chinese and Western medicine | 4 | 5.71 |
| Public health management | 3 | 4.29 |
| Clinical pharmacy | 3 | 4.29 |
| Others <sup>b</sup> | 5 | 7.14 |

*Note:* a: including internal medicine, surgery, and pediatrics;
b: including preventive medicine, rehabilitation therapeutics, medical imaging, medical imaging technology, and
medical laboratory science.

### Statistical descriptive analysis of condition and outcome variables

Appendix 6 details the coding result of the variables. Analysis of the IFQ questionnaire for the condition variables revealed a skewed distribution, leading to the adoption of medians (*M*) and interquartile ranges (*IQR*) for statistical description (Appendix 7). Among the 12 institutions, the influence levels of each condition factor were generally rated as "3=fairly influential", with median scores varying between 2.52 to 3.31 and median scores for outcome variables at 105.20 (101.23 to 107.33).

Furthermore, the WRQ-CN results, segmented by dimension, indicated medians and *IQR* as follows: Context at 34.10 (33.28 to 36.25), Change valence at 18.83 (17.92 to 19.99), Informational assessment at 18.71 (18.25 to 19.10), Change commitment median score 17.67 (16.66 to 18.75), and Change efficacy at 14.43 (13.71 to 15.29).

### Correlation analysis between the condition and outcome variables

The results from *Spearman* correlation analysis showed that under the *Coherence* domain (Appendix 8a), there was a moderate correlation between the value of *Specific tasks and responsibilities* and the WRQ-CN value, with an *r*-value of s 0.393 (*p* = 0.206). The correlation between *Key participants* and the WRQ-CN value had a moderate *r-*value of 0.316 (*p* = 0.317) (Appendix 8b). The *r* values for the other correlations were ranged from 0.025 to 0.294.

### Exploration of pathways to a high-level of ORC Truth table analysis

We chose four conditions for *Coherence* and seven for Cognitive Participation in conducting the truth table and configuration analysis, respectively (The variable calibration and necessity analysis results were presented in Appendix 9). The distribution of the raw consistency shows variations ranging from 0.57-0.94 and 0.49-0.93, respectively, in two domains. The consistency threshold value set for this study was 0.8. Thus, in Coherence and Cognitive Participation, the configuration of institutions A, E, F, G, and H showed a good correlation for the outcome variable, which means there was a correlation between the configuration of these institutions and high levels of ORC (Table 2).

**Table 2.**
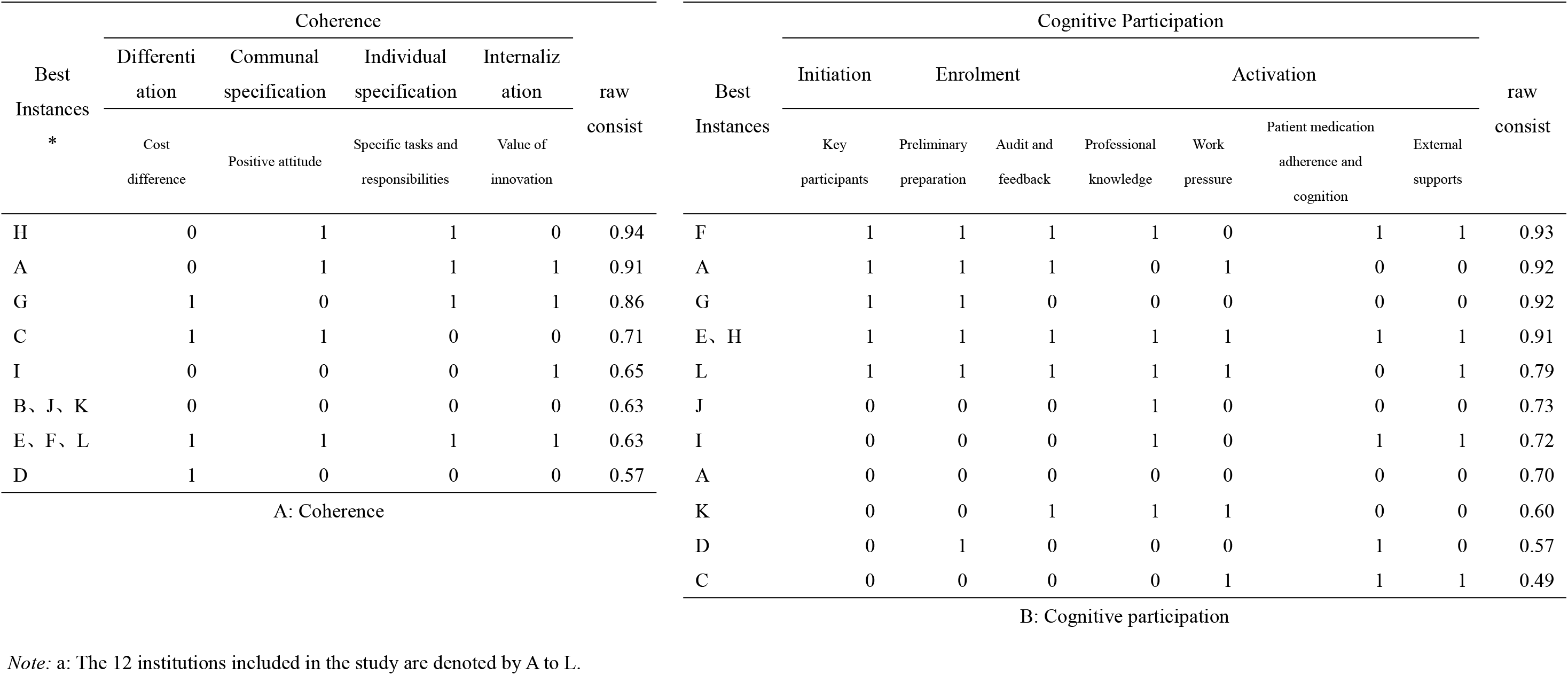
The Qualitative Comparative Analysis results (n = 12 institutions)

### Configuration analysis

Under the standard analysis, The solution consistency values of different configurations were all greater than 0.8, which means there were strong correlations between different configurations and high-level ORC. The solution coverage values were 0.49 and 0.60 in *Coherence* (Figure 1A) and *Cognitive Participation* (Figure 1B), respectively, presenting the configurations that collectively explained 49% and 60% of the high-level ORC cases.

**Figure 1.**
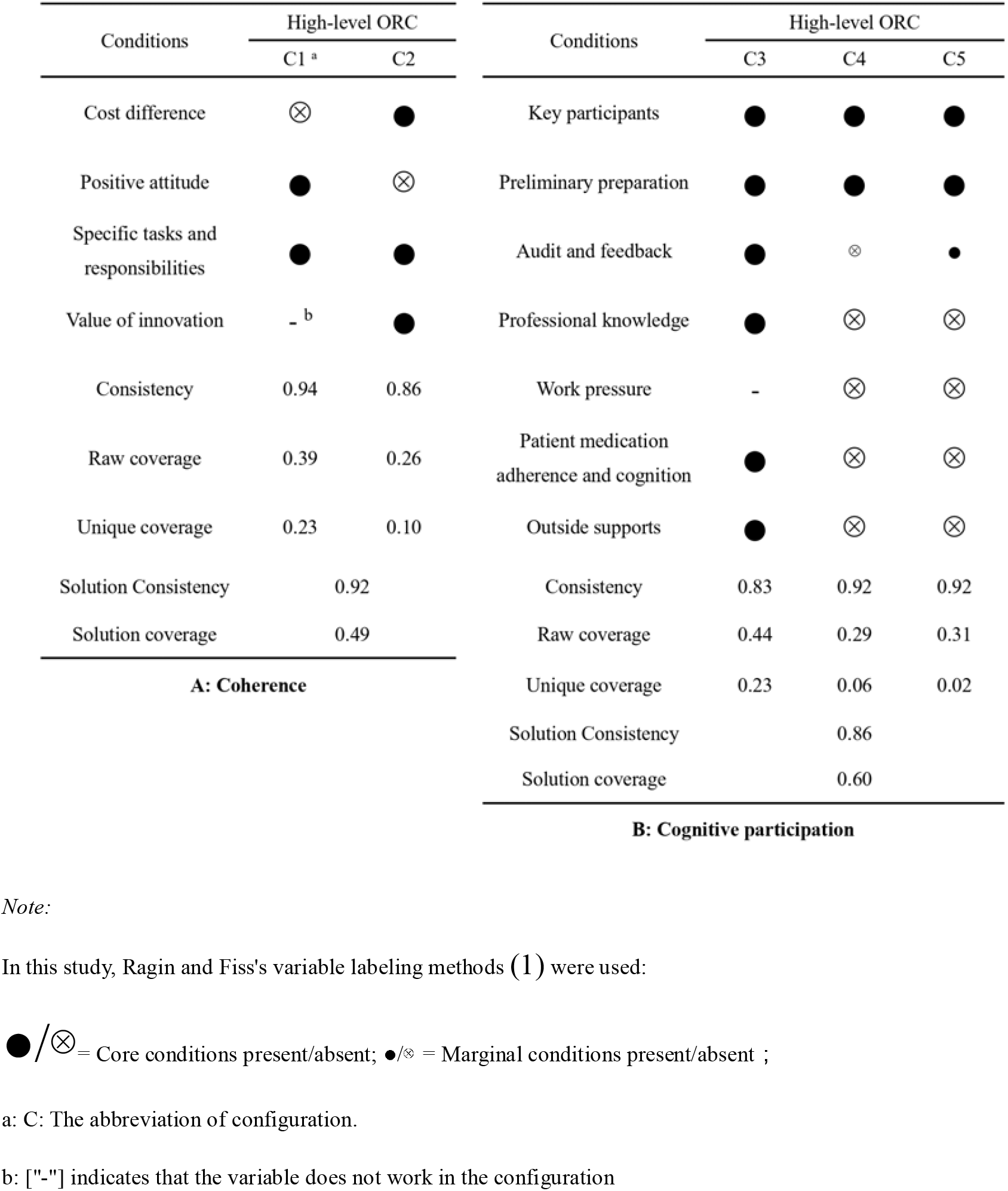
Configuration analysis

In the *Coherence*, two configurations could help the organization achieve high-level ORC. First, the type C1 (i.e., *Community-perceived type) presented t*he interviewees were willing to participate (i.e., Positive attitude) in SMA actively, understood the SMA service *Specific tasks and responsibilities* when implementing SMA would help them achieve a higher level of ORC. Second, the type C2 (i.e., *Well-understanding type*) presented the interviewees could tell the *Cost differences* (time, human resources, and money) between the SMA and formal service, understood the SMA *Specific tasks and responsibilities* when implementing SMA, and agreed with the value of implementing the SMA (i.e., the value of innovation) would help them achieve a higher level of ORC.

In *Cognitive participation*, configurations could help the organization achieve a high-level ORC. First, the type C3 (i.e., *Everything-was-ready type*) presented prior to initiating SMA to recruit patient participants and provide service, the presence of the following condition variables can significantly enhance the ORC: 1) *Key participants* led the SMA pilot trial implementation; 2) the staff work together to prepare the consultation room and the needed materials, reorganize the cooperation model between each other (i.e., *Preliminary preparation*); 3) carry out *Audit and feedback;* 4)consolidate *Professional knowledge;* 5) *Patients’ medication adherence and cognition* on diabetes were high; 6) and availability of a variety of *external support*, including support from higher level administrative department and research teams guidance. Second, type C4 (i.e., the driving force was needed type) presented *Key participants*, and *Preliminary preparation* was the core of the configuration, which both improved the ORC and achieved a higher level. Third, type C5 (i.e., *type C4 plus*) was very similar to C4, and the difference was that in addition to the *Key participants* and the *Preliminary preparation,* the *Audit and feedback* played a role as a marginal condition to improve the ORC.

#### Pathway Analysis

*Coherence* and *Cognitive Participation* were distinct domains, but they can be seen as consecutive linked phases in the mindset shift preceding the implementing innovation^25^. To achieve high-level ORC, we integrated the previously discovered five configurations of these two stages. Therefore, we identified and selected key conditions, both core and marginal conditions, alongside variables demonstrating moderate quantitative correlation, including *Specific tasks and responsibilities, Key participants, Preliminary preparation*, and *Audit and feedback.* Further analysis by QCA indicated two types of configurations (i.e., combination conditions) to achieve high-level ORC (Figure 2). The solution consistency was 0.94, which means that the combination of conditions predicted the outcome in 94% of the cases. This was a relatively high level of consistency and indicates a significant correlation between the conditions and the outcome^46^. The coverage was 0.39, which presented the configurations collectively explaining 39% of the high-level ORC cases.

**Figure 2.**
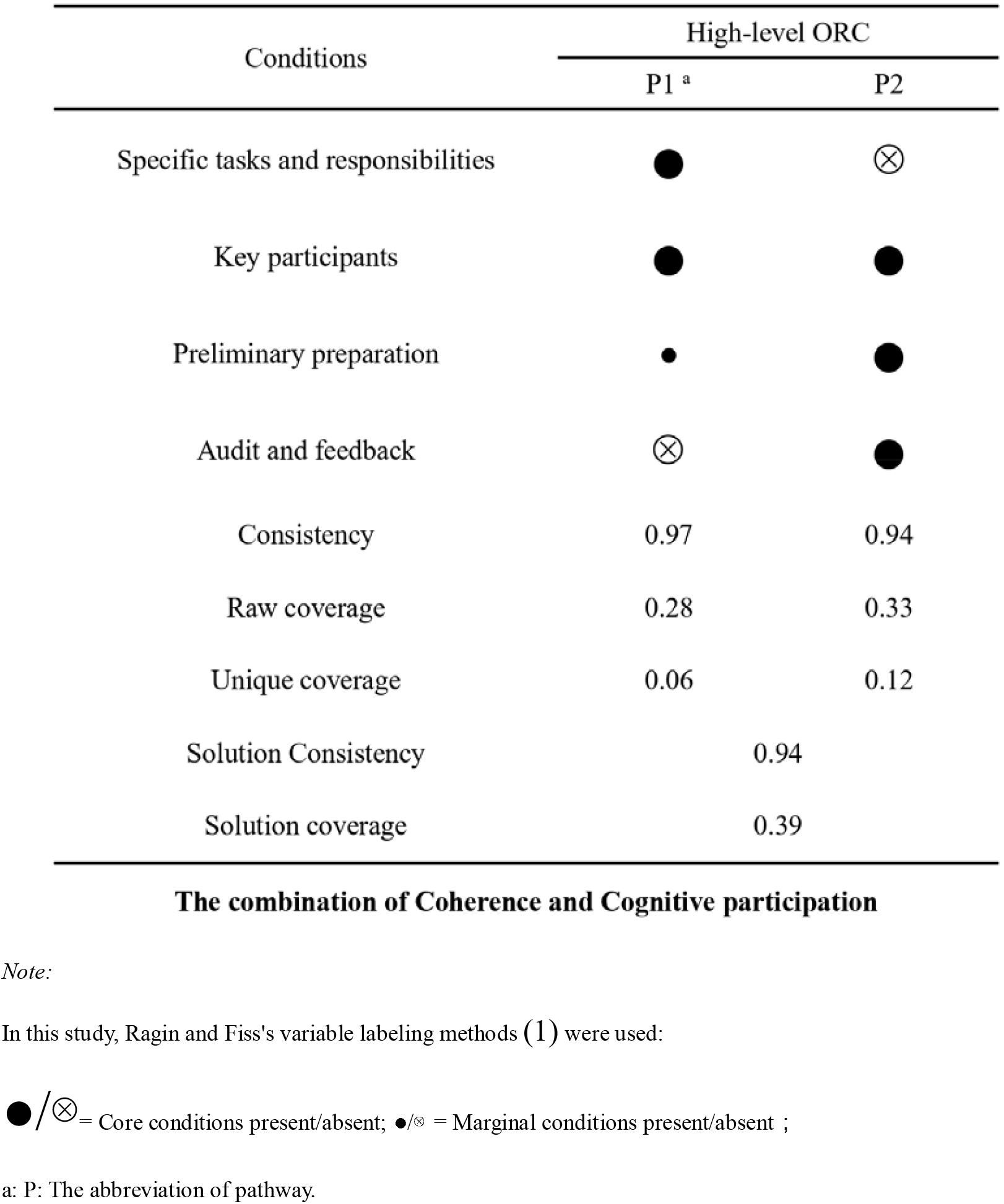
Pathway analysis

The two configurations presented two pathways consistent with the NPT theory. In P1, when participants were aware of their *Specific tasks and responsibilities* within the *SMART* pilot trial (i.e., belonging to the *Coherence* domain), the *Key participants* enhanced the process of *Preliminary preparation* of the pilot trial (i.e., belonging to the *Cognitive Participation* domain), the high-level ORC would easily be achieved. Conversely, in situations where the participants were unclear about their *Specific tasks and responsibilities*, but the *Key participants* assumed additional responsibilities to ensure thorough *Preliminary preparation* and provide *Audit and feedback*, the high-level ORC would also be achieved (P2). The pathway of the sequential and concurrent condition to achieve high-level ORC was presented in Figure 3.

**Figure 3.**
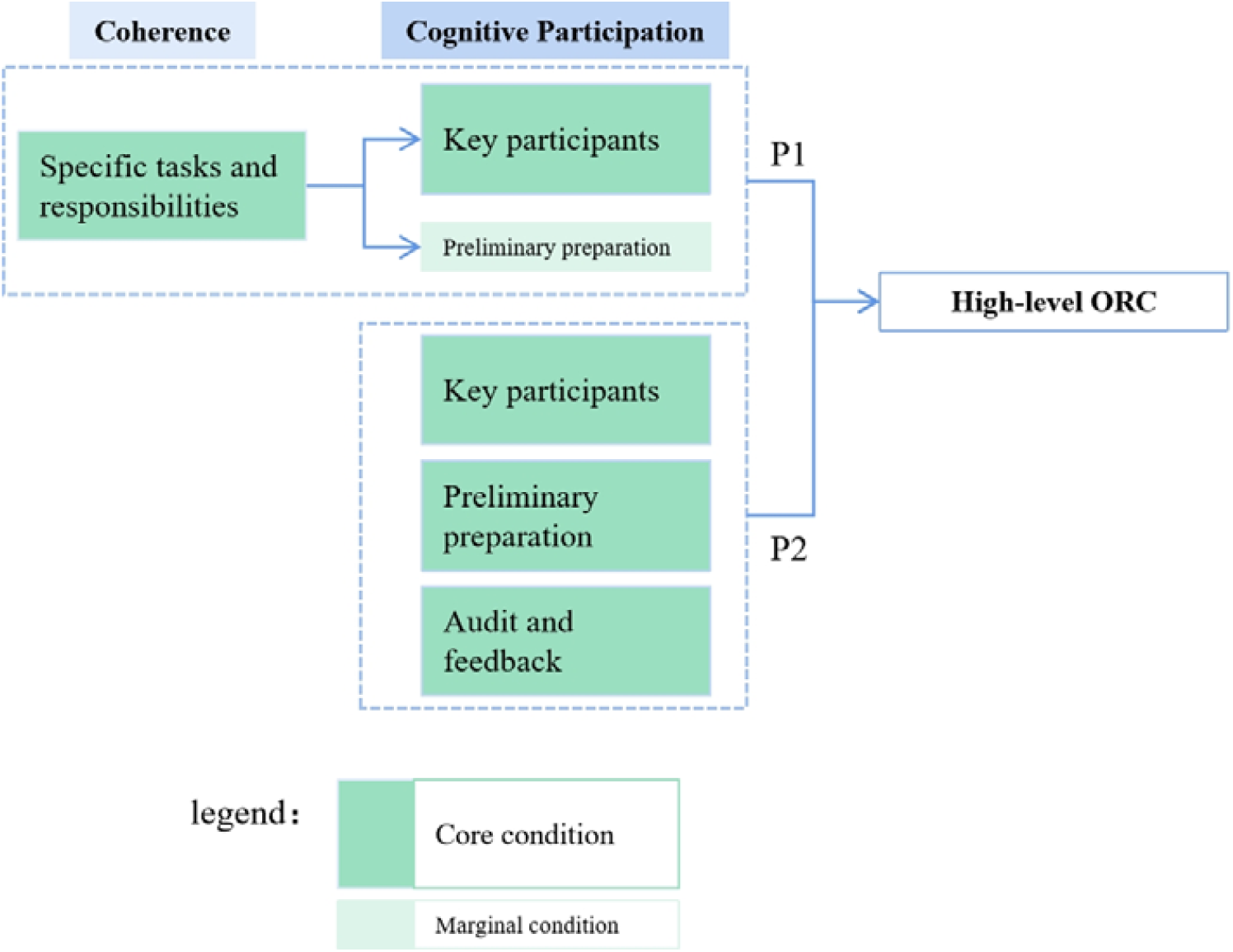
The pathways to achieve high-level ORC

#### Robustness test

We conducted two kinds of robustness tests by adjusting the consistency threshold from 0.80 to 0.85 and added the variable of *Valid contribution for the innovation* as one of the condition variables related to the ORC. It was found that whether we adjusted the consistency threshold or added the variables related to the outcome, the key parameters, including the variables in different configurations, the solution consistency, and the solution coverage were only slightly different from the initial primary result, and no substantial changes occurred (Appendix 10).

## Discussion

Our study investigated factors (i.e., condition variables) affecting the ORC within *Coherence* and *Cognitive Participation* and identifying five concurrent configurations of achieving high-level ORC, which were C1 - *Community-perceived*, C2 - *Well-understanding*, C3 - *Everything-was-ready*, C4 - *Driving force was needed* and C5-*Supplement of C4*. Given the interconnectedness of *Coherence* and *Cognitive Participation* in the implementation innovation^22^ ^23^, we integrated prevalent factors from these configurations for further QCA. This revealed two concurrent pathways to high-level ORC, highlighting the critical role of *Key participants*.

### Different types of configurations for achieving high-level ORC

In the *Coherence* domain, the Type C1 - *Community-perceived* showed that when the service providers kept a *Positive attitude* toward innovation and knew the *Specific tasks and responsibilities,* the ORC would achieve a higher level. Individuals promoted the implementation of innovation and motivated the collective responsibility of the others in their organization. Studies have shown that individual and organizational consensus on interventions was closely related to work efficiency^47,48^. Thus, participant consensus at the *Coherence* could build a team-based implementation atmosphere that improves ORC, extends innovation’s effectiveness, and promotes its success.

Type C2 – *Well-understanding* showed that the *Cost difference*, *Specific tasks and responsibilities*, and *value of innovation* played a core role in bringing an organization’s ORC to its desired status. In our study, institution G resented completely the same pattern as C2. Staff in Institution G were active and very willing to participate in our study, so they presented an active attitude after the launching and training session. When the staff understood and valued the innovation, the *Change valence* was high, and a high-level ORC would appear^49^.

In types C1 and C2 under the *Coherence* domain, understanding the *Specific tasks and responsibilities* played a core role in getting a high-level ORC. Meanwhile, correlation analyses showed that the indicator was moderately correlated with the outcome variable. These two similar results indicated that the individual’s ideological change drove other staff to change their thoughts and behaviors towards the SMA to reach a high-level ORC and would facilitate the implementation of the SMA in the next step.

In the Cognitive Participation domain, the type C3 - Everything-was-ready showed that when there were Key participants, good Preliminary preparation, regular Audit and feedback, equipped Professional knowledge, high level of Patient medication adherence and cognition, and the available Outside (environmental) support to the organization, which included the most number of condition variables, the ORC will achieve a good condition. Although previous studies have found a positive correlation between Work pressure and readiness for innovation^50^, we did not find the same effect of participants’ Work pressure on ORC. In our study, Work pressure would no longer affect the ORC status when the variables mentioned above were included. The front-line workers in this study were already under relatively high stress before participating in the *SMART study*. When the staff was motivated to work together with the researcher to implement the *SMART study* in the training session and find an optimal type of SMA, the innovation could release their work pressure in daily service. So, they were eager to reduce their workload through the SMA innovation. In this situation, the Work pressure of the SMA innovation would no longer affect their preparation for implementing the SMA pilot trial.

In type C4 – *A driving force was needed*. If the institutions had *Key participants* to drive the SMA innovation and conduct *Preliminary preparation* well, the organization would achieve a high-level ORC status. This finding was consistent with previous studies that showed that strengthening leadership and innovation awareness through training before implementing innovation could improve the ORC^51^. The contribution of *Key participants* and *Preliminary preparation* were dominant for high-level ORC in this type. In our study, Institution G made sufficient *preliminary preparations* (consultation room layout, education materials, staff cooperation, etc.), and its *Key participant* was the institution’s administrator, who also acted as a clinician and played a coordinating role in the implementation process. With these two conditions, the ORC level achieved a high level.

The type C5 – *type C4 plus* contained conditions similar to C4 and added *Audit and feedback. Audit and feedback* effectively improved the successful implementation of innovation^52^ ^53^. The high-level ORC would also be achieved if the administrators provided *audits and feedback* besides the contribution of *Key participants and preliminary preparation* to the staff. So, we could regard Type C5 as a Type C4 plus. In our study, Institution A presented the same pattern as C5. In Institution A, the administrator and the clinician both played the role *of Key participants, leading the Preliminary preparation and providing Audits and feedback*.

Sometimes, the administrator initiates an innovation without involving the staff’s opinion. The preparation process and mind readiness would lag behind other institutions where staff and administrators were eager for innovation^54^. Although work pressure and regular Audits and feedback can motivate employees to engage more in innovation and successfully improve the ORC, this was not the best solution to improve readiness^55^. Because participants’ engagement was critical to arousing their awareness of conducting innovation^56^ ^57^, involving the staff in finding the benefits of implementing innovation, they would start preparing and achieving a high level of innovation.

### The pathway to achieving high-level ORC

In implementation research, *Coherence* and *Cognitive Participation* were sequential and interrelated phases that facilitate a mindset shift before implementing innovation . Utilizing NPT-guided interviews and data analysis, we construct an explanatory result. The results indicated that the combined essential conditions could further present two consecutive pathways that can achieve high-level ORC concurrently. This conclusion was drawn from the correlation analysis and QCA statistics and, corroborated by empirical evidence, and can be consistent with the NPT-based dynamic steps.

In our study, Institutions G and A presented a similar pattern to pathways P1 and P2 separately. Following the training, staff members in Institution G quickly grasped their *Specific tasks and responsibilities*. The *Key participant* (i.e., their administrator) expressed that he would take responsibility for preparing the pilot trial. Conversely, Institution A’s staff initially lacked a clear understanding of their *Specific tasks and responsibilities*. However, their administrator and the clinician would prepare the pilot trial in their institution, including setting up the consultation rooms and necessary materials. The administrator and the clinician provided *Audits and feedback* to each other. Finally, they also achieved a high-level ORC. And then, the two institutions implemented the pilot trial well in our study. Similar to how *Audit and feedback* have been validated as an effective strategy for enhancing implementation innovation^58^. This situation could also be found in other studies^59^ ^60^. The *Key participants* (opinion leaders, internal implementation leaders, champions, outside innovation drivers, etc.) leading the innovation and involving participants conducting *Preliminary preparation(s)* would be essential condition variables in facilitating the innovation’s readiness.

Our study found a moderate correlation between *Key participants’s* involvement and ORC. Key participants, typically the administrators, played a crucial role in spearheading innovation, motivating and engaging staff in the process. This leadership facilitated a collective understanding of roles and contributed to effective preliminary preparation. We documented all these conditions before assessing their ORC status. Therefore, the influence of these conditions on the ORC would be viewed in a longitudinal relationship. We proposed that there might be causal relationships existed between these conditions and ORC. However, the precise mechanisms underliying this relationship still need to be rigorously tested in randomized controlled trials. Furthermore, based on these relations, future studies can explore implementation strategies using the Expert Recommendation for Implementing Change (ERIC)^61^, and validate their effectivenesss in improving the readiness in real-world applications.

### Limitations

This study has several limitations impacting its external validity and interpretative scope within the field of implementation science. First, we chose 12 institutions by purposive sampling to present the diversity in their geographic location, medical resources, chronic disease care service capacity, and human resources. However, the sample size was relatively limited, which may constrain the generalizability of our findings. Second, we only assessed ORC prior to innovation implementation, ignoring its dynamic nature and the evolving engagement of participants^62–64^, Future research will examine the changes in ORC and its determinants. Third, the *SMART* pilot trial was a three-factor, two-level factorial design that could predict the main effects and interaction between the condition variables in the SMA components^14^, leading to the existence of different intervention types, which may indirectly result in uneven levels of difficulties of prepare the SMA service. However, we assume that this could only affect the visible preparation (i.e., consultation room preparation, health management preparation), other than the readiness in their mind.

## Conclusions

The study revealed a constellation of conditions and two dynamic pathways that impacted the ORC, aligning with the consecutive and interactive steps proposed by the NPT. Though the processes of coherence and cognitive participation, engaging the key participants, through comprehensive preliminary preparation, and a providing audit and feedback to other participants could boost ORC.

The research highlighted the intricate and dynamic nature of achieving organizational readiness, pinpointing crucial conditions that facilitate the process. Employing the QCA approach allowed for a comprehensive understanding of how these conditions interact with ORC status. Such insights could guide others toward more efficient and scaled-up implementation of health innovations.

## Abbreviations

CCM: Chronic Care Model
CHCs: Health Service Centers
DM: Diabetes mellitus
EBP: Evidence-Based Practice
ERIC: Expert Recommendation for Implementing Change
fsQCA: fuzzy-sets QCA
IFQ questionnaire: Influencing Factor Quantitative questionnaire
IQR: interquartile ranges
M: medians
MOST: Multiphase Optimization Strategy
MMARS: Mixed Methods Article Reporting Standards
NEPHS Program: National Essential Public Health Services Program
NPT: Normalization Process Theory
ORC: Organizational Readiness for Change
ORC Theory: Weiner’s theory of Organizational Readiness for Change
PWD: patient with diabetes
QL: lower quartile,
QU: upper quartile
QCA: Qualitative Comparative Analysis
REDCap: Research Electronic Data Capture platform
SMA: Shared Medical Appointment
SMART: Shared Medical Appointment for diabetes
STROBE: Strengthening Reporting of Observational Studies in Epidemiology
TA: Thematic Analysis
T2DM: type 2 diabetes Mellitus
THCs: Township Health Centers
WRQ: Workplace Readiness Questionnaire
WRQ-CN: Workplace Readiness Questionnaire - Chinese

## Authors’ contributions

YC conceptualized this study and set up a core implementation science team to analyze the data. This team included the two leaders (YC and DRX), a QCA methodologist (LL), two research assistants (WY and RM), and Four on-site data collection coordinators (JC, TY, LhL, and LH). All had intimate knowledge of the study protocol and previous quantitative and qualitative data collection and analysis experience, and all conducted and/or analyzed the interviews. WY collected or maintained all data and wrote the first draft of the manuscript. YC refined this manuscript. All authors read and approved the final manuscript.

## Ethics approval and consent to participate

This study was conducted under the National Natural Science Foundation of China (NNSFC).

Shared Ethics Approval Document with NNSFC as a sub-theme of its (The Ethics Committee of Guizhou Medical University Ethics Approval Document 2023 (4)]: https://kdocs.cn/l/cjoKKtlCBop4). The study was conducted after the participants were informed of the cautions and had signed the informed consent form. Meanwhile, we audio-recorded the interviews.

## Trial registration

ChiCTR2300069904

## Availability of data and materials

The study does not share the original data. If there are any special needs, the datasets used and analyzed during the study are available from the corresponding author or the first author’s request.

## Acknowledgments

We thank Peggy A. Hannon, the developer of the WRQ, for authorizing us to use the scale. We thank all the participants of the CHC/THC of Bozhou District, Zunyi City, and the CHC/THC of Bijiang District, Tongren City. Without your support, we could not have completed the study.

