## Supplemental Data 1 for "Factors Influencing the Implementing Readiness of Shared Medical Appointments in China’s Primary Healthcare Institutions: A Mixed-Method Study Utilizing Qualitative Comparative Analysis"

### Additional files

#### Appendix 1 Operationalization of NPT in this study

| 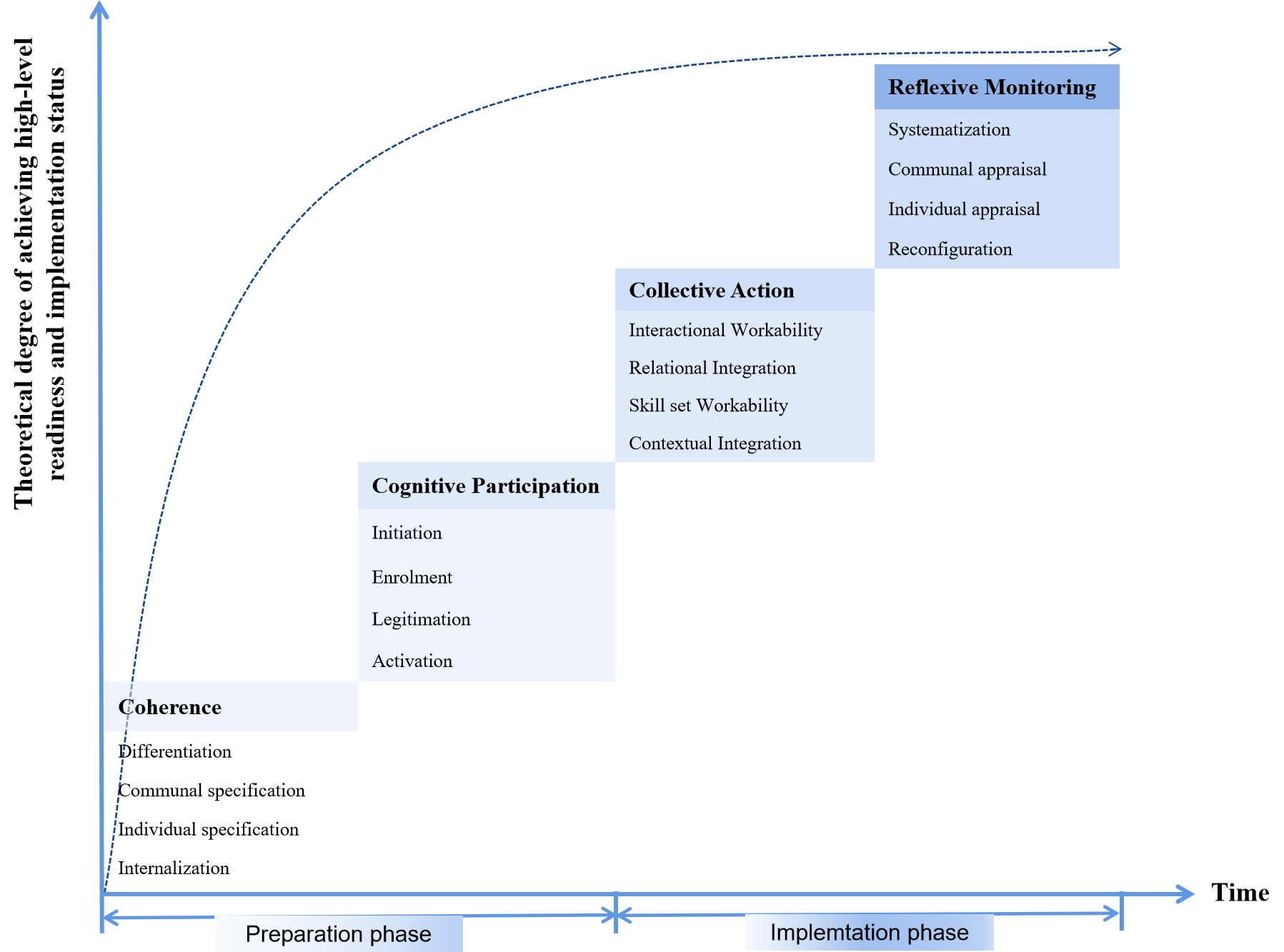 | 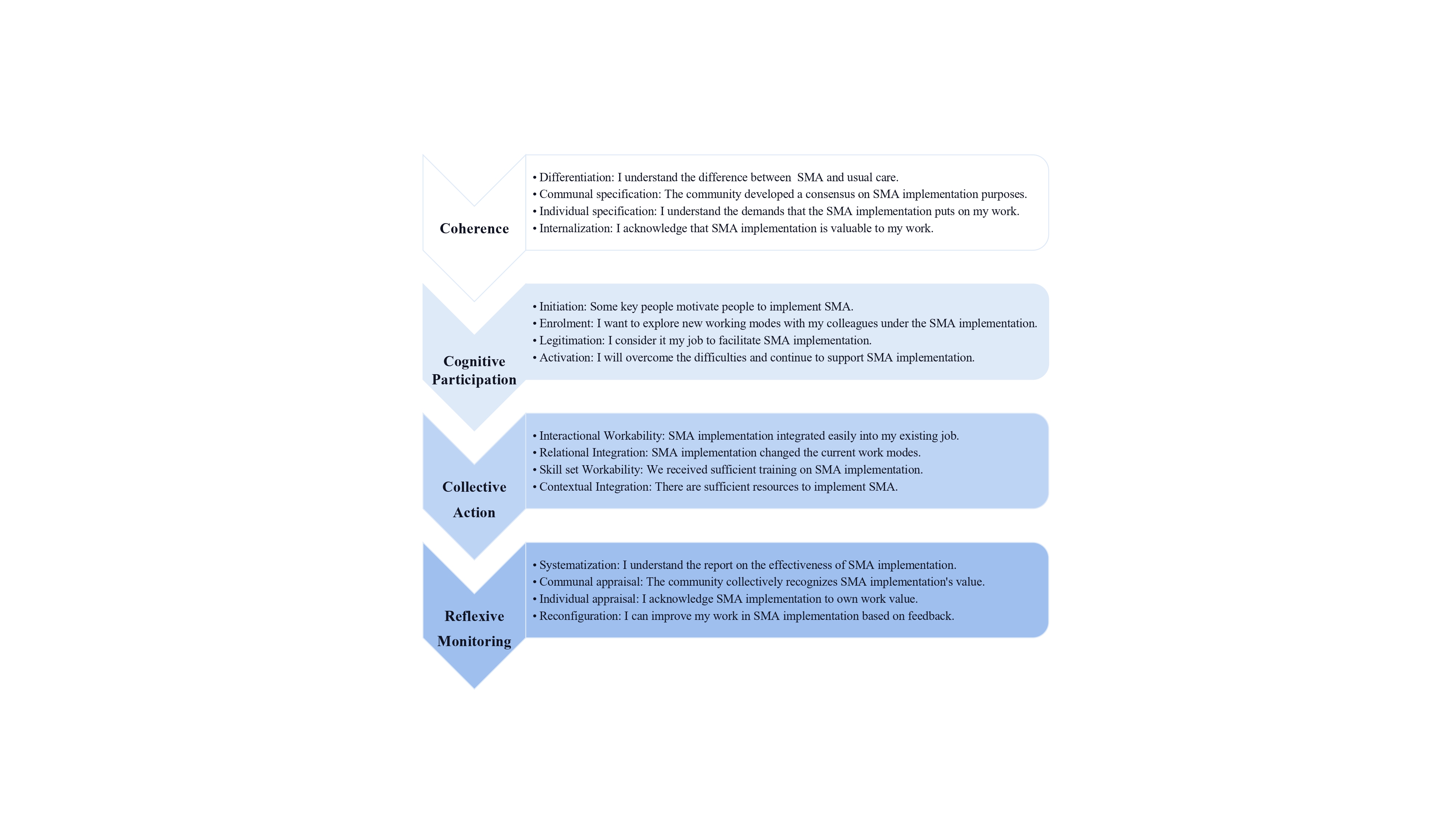 |
| --- | --- |
| (A) NPT-based hypothetical sequences of achieving readiness and implementation | (B) NPT-based interview outline |

#### Appendix 2 The WRQ-CN and WRQ Detailed Version

WRQ

Context

Q1. The senior leaders are willing to try new things.

1Never	2Rarely	3Sometimes	4Often	5Always

Q2. The senior leaders seek ways to improve the work climate.

1Never	2Rarely	3Sometimes	4Often	5Always

Q3. The senior leaders reward creativity and innovation in the worksite.

1Never	2Rarely	3Sometimes	4Often	5Always

Q4. The senior leaders promote team building to solve worksite problems.

1Never	2Rarely	3Sometimes	4Often	5Always

Q5. The managers seek ways to improve the work climate.

1Never	2Rarely	3Sometimes	4Often	5Always

Q6. The managers encourage employees to participate in programs.

1Never	2Rarely	3Sometimes	4Often	5Always

Q7. Opinion leaders are willing to try new things.

1Never	2Rarely	3Sometimes	4Often	5Always

Q8. Opinion leaders seek ways to improve the work climate.

1Never	2Rarely	3Sometimes	4Often	5Always

Q9. When we want to try something new we have the training resources to do it.

1Never	2Rarely	3Sometimes	4Often	5Always

Q10. When we introduce a new program or change we measure its success by asking employees to fill out a survey about the program.

1Never	2Rarely	3Sometimes	4Often	5Always

Change valence

Q11. We measure employee participation in wellness programs.

1Yes	2No

Q12. We measure employee satisfaction with our wellness programs.

1Yes	2No

Q13. We seek information about wellness from brokers or others outside our organization.

1Yes	2No

Q14. Our organization has established, written wellness goals.

1Yes	2No

Q15. Wellness programs would improve employee health in my organization. IF RESPONDENT ASKS IF WE WANT THEIR PERSONAL OPINION OR THAT OF THE ORGANIZATION, SAY: "We would like to know your personal opinion".

1Strongly Disagree	2Disagree	3Neutral	4Agree	5Strongly Agree

Q16. Wellness programs reduce employers' health care costs.

1Strongly Disagree	2Disagree	3Neutral	4Agree	5Strongly Agree

Q17. Wellness programs help companies recruit and retain employees.

1Strongly Disagree	2Disagree	3Neutral	4Agree	5Strongly Agree

Q18. Wellness programs are a good use of financial resources.

1Strongly Disagree	2Disagree	3Neutral	4Agree	5Strongly Agree

Informational assessment

Q19. Most employees could take time at work to participate in wellness programs.

1Strongly Disagree	2Disagree	3Neutral	4Agree	5Strongly Agree

Q20. Senior leaders would dedicate financial resources to wellness programs.

1Strongly Disagree	2Disagree	3Neutral	4Agree	5Strongly Agree

Q21. Senior leaders would dedicate staff time to planning wellness programs.

1Strongly Disagree	2Disagree	3Neutral	4Agree	5Strongly Agree

Q22. We have one or more employees who are wellness champions.

1Strongly Disagree	2Disagree	3Neutral	4Agree	5Strongly Agree

Q23. We have one or more senior leaders or managers who are wellness champions.

1Strongly Disagree	2Disagree	3Neutral	4Agree	5Strongly Agree

Change commitment

Q24. Our senior leaders are committed to improving our (starting a) wellness program.

1Strongly Disagree	2Disagree	3Neutral	4Agree	5Strongly Agree

Q25. Our opinion leaders are committed to improving our (starting a) wellness program.

1Strongly Disagree	2Disagree	3Neutral	4Agree	5Strongly Agree

Q26. We are motivated to improve our (implement a) wellness program.

1Strongly Disagree	2Disagree	3Neutral	4Agree	5Strongly Agree

Q27. We need to improve our (start a) wellness program within the next year.

1Strongly Disagree	2Disagree	3Neutral	4Agree	5Strongly Agree

Q28. We have the skills and expertise to expand our (implement a) wellness program.

1Strongly Disagree	2Disagree	3Neutral	4Agree	5Strongly Agree

Change efficacy

Q29. We have enough financial resources to support a wellness program.

1Strongly Disagree	2Disagree	3Neutral	4Agree	5Strongly Agree

Q30. We can (could) manage the politics of implementing a wellness program. IF RESPONDENT IS UNCERTAIN, SAY: "This means that you could manage the different activities, relationships, and personalities involved in implementing a wellness program at your organization.

1Strongly Disagree	2Disagree	3Neutral	4Agree	5Strongly Agree

Q31. We can (could) get people to participate in our wellness program.

1Strongly Disagree	2Disagree	3Neutral	4Agree	5Strongly Agree

Q32. How much time do you think you could spend each week on managing a wellness program?

1Strongly Disagree	2Disagree	3Neutral	4Agree	5Strongly Agree

#### Appendix 3 Pilot organization coverage areas

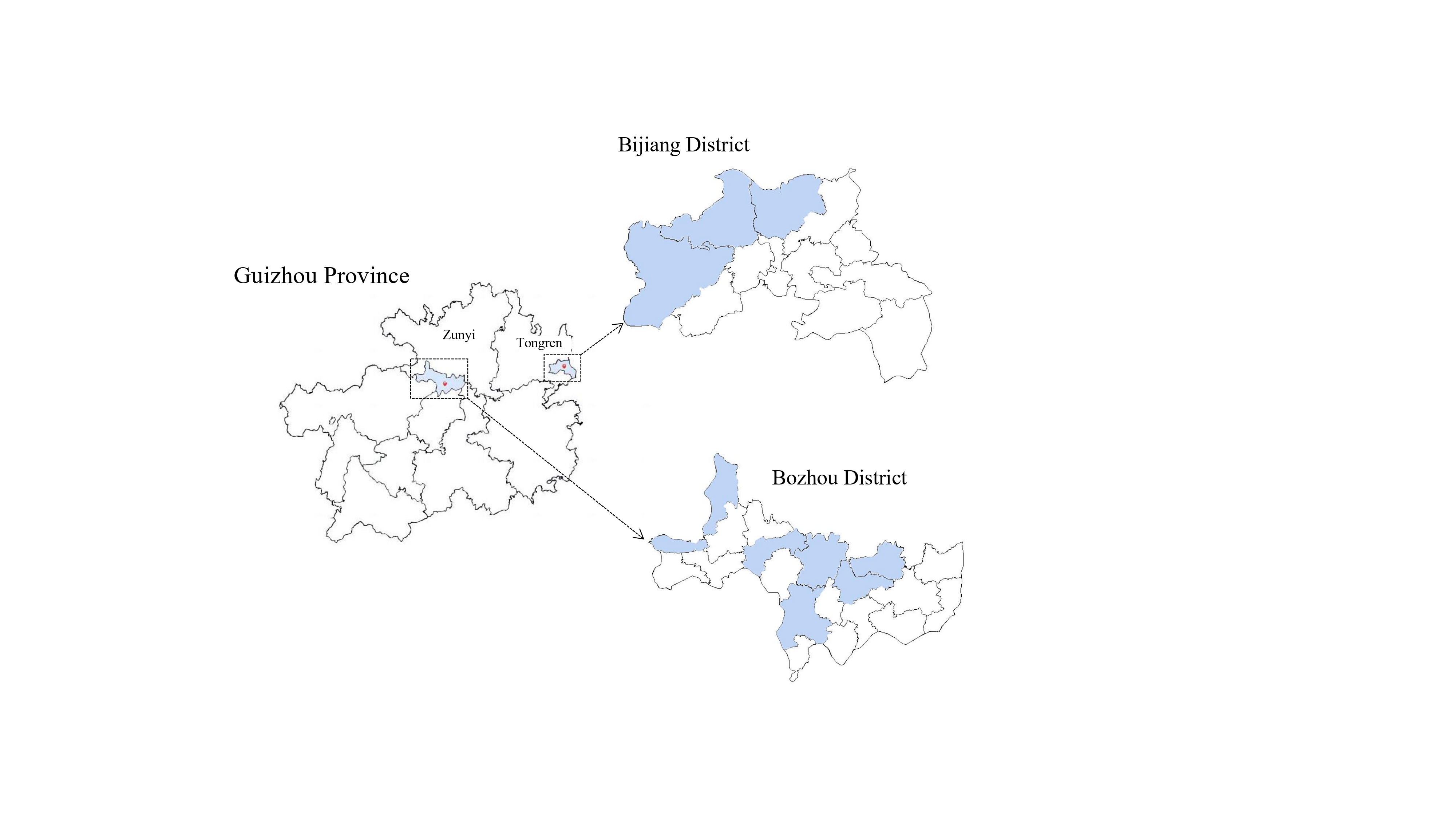

#### Appendix 4 Qualitative Comparative Analysis

QCA was a qualitative analysis method based on Boolean algebra and Set-Theory for case comparisons, exploring complex social phenomena by seeking causal relationships between variables and identifying configurations that can achieve desired results (1). It was commonly used in sociology, management, and healthcare (2-5). QCA can identify the necessary conditions and conditional configuration sufficiency of a high-level of ORC, help answer questions about multiple concurrent causality and causal non-symmetry, better match the interdependence and causal complexity of implementing innovation in complex healthcare (6,7), compensate for the limitations of quantitative analysis that departs from causal complexity (8), and better result in a realistic model reflecting the complexity of healthcare to maximize and normalize the benefit of the innovation (9).

It was necessary to determine the degree of membership of various cases to a set in QCA based on the fuzzy-set membership scores (0-1) of the variables, including two qualitative states: 1 = full membership and 0 = full nonmembership; 0.5 to 1 for strong membership; and 0 to 0.5 for weak membership. 0.5 is the qualitative locus point, the maximum fuzziness for assessing whether a case is a membership or nonmembership (6).

After Standard Analyses in QCA, three scenario solutions can be obtained (6). The Complex Solution consists of a combination of multiple conditions, each of which contributes to the result; the Intermediate Solution consists of a combination of a few conditions that still explains the result; and the Parsimonious Solution consists of a combination of only one or very few conditions that was sufficient to explain the result (10-12).

QCA truth tables reported on raw consist, indicating for each truth table row which shows results in the proportion of cases. The following values are reported for conditional configurations that generate high readiness before innovation implementation. ①Consistency: measures the extent to which each configuration was a subset of the result set; ②Raw coverage is the proportion of cases that match the configuration; ③Unique coverage means the proportion of cases that uniquely fit the configuration but not the others; ④Solution consistency means assessing the fitness of a model by comparing the frequency of different configurations; ⑤Solution coverage is the proportion of cases covered by all configurations (2,10,13-15). The study interpreted configurations with consistency thresholds ≥ 0.8; coverage does not currently have an acceptable minimum threshold, and higher values are usually considered to have high experience relevance or significance but need to be interpreted by the researcher in a context-specific (16).

#### Appendix 5 Study design and data analysis process

**① Interview text analysis and coding.** All audio recordings were transcribed at the end of the interviews to support the credibility of the research findings. By constantly comparing and categorizing data, multi-level and multi-dimensional observations were developed. First, the interview texts were transcribed to support the reliability of the results and analyzed using Nvivo 12.0 based on NPT using TA (1). Then, a variable set of influencing condition variables was developed based on the coding of the NPT, and the variable set was used to develop the IFQ questionnaire, which was utilized to conduct a Full-Sample Survey to form the condition variables for the subsequent analyses. The researcher interpreted the results based on practical experiences, dug deeper, and found the essential features of the variables corresponding with the theory.

1. **Statistical characterization and *Spearman* correlation analysis.** Summarized the results of the IFQ questionnaire and Readiness Scale survey, used Jamovi 2.3 to calculate their statistical distributions and analyzed the variables' correlations based on the characteristics of the distributions. When the correlation coefficient *r* ≥ 0.3, the variables were moderately correlated and above (2), and their correlation coefficients were plotted.
2. **Variables calibration and membership** **calculation.** Calibration is the process of assigning a membership score to a variable (3). Membership is the degree of association or similarity between a variable and a particular attribute. It is usually indicated by using a value between 0 and 1, where 0 indicates full nonmembership, and 1 indicates full membership (4). In this study, based on the numerical characteristics of the variables and Ordanini's study (5), the variables were calibrated using the direct calibration method to determine the casual and outcome variables calibration points, i.e., Jamovi 2.0 was utilized to calculate the corresponding values of each variable at the 90th, 50th, and 10th percentiles based on the percentile method (3,4,6). The corresponding values were brought into the fsQCA 3.0 software calibrate (x,n1,n2,n3) (3,7). To avoid ignoring crossover points affecting the results to increase the feasibility and completeness of the analysis (8). Eventually, all cases were categorized into two calibrated standard sets: fully affiliated and fully unaffiliated.

In QCA, the calibrate(x,n1,n2,n3) is a method used to convert raw data into membership. It is based on the principles of fuzzy set theory, which means that the membership of each case to a set can be any value between 0 and 1, not just 0 or 1 (4). This can better reflect the continuity and variability of the case.

Calibrate(x,n1,n2,n3) calculates the membership as follows (4):

Equation 1:
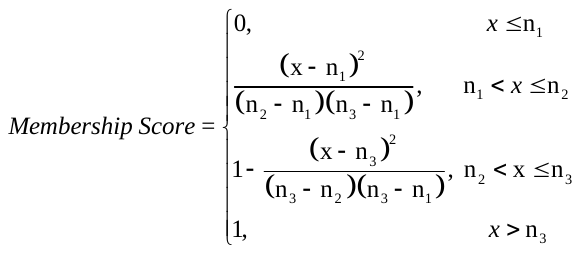

Where x is the value of the original data after calibration, n1 is the threshold for full nonmembership, n3 is the threshold for full membership, and n2 is the crossover point. A crossover point is a point with a membership degree of 0.5, i.e., the cases have the same degree of membership and nonmembership for the set.

1. **Necessity and coverage analysis.** The necessity test is to assess whether a variable constitutes a necessary condition for the outcome, i.e., the outcome would not be possible in the absence of the variable, and a necessary condition is a subset of the outcome, which is present in all cases with the outcome (4). The principle is to determine whether the condition is a subset of the outcome by comparing the membership of the condition and the outcome. If the membership of the condition is less than or equal to the membership of the outcome, then the condition is a necessary condition for the outcome, and conversely, the condition is not a necessary condition for the outcome (6). The consistency is an important indicator to assess a necessary condition (9). It refers to the degree of subset relationship between the condition and the result, with a lower limit of typically 0.9 (3,6). If consistency < 0.9, the condition is not necessary, i.e., the variable is not necessarily present in the configuration.

The consistency Equation is as follows (3,10):

Equation 2:
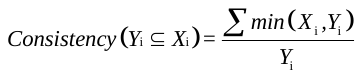

For the assessment of sufficiency, coverage is mainly used, which is calculated using the following equation:

Equation 3:
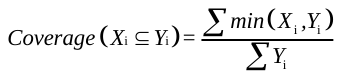

Where *X* is the condition variable, *Y* is the outcome variable, and *X*_i_ and *Y*_i_ are the membership of the condition and outcome of the i case, i.e., the sum of the membership of the condition variable divided by the membership of that variable. For cases that are inconsistent with the subset relationship, the part of the configuration where the membership is consistent with the outcome can be used to achieve this. min(*X*_i_, *Y*_i_) is the minimum of the two when *X* is compared with *Y*, i.e., when *X* value is greater than *Y*, the value of *Y* is brought into the calculation.

1. **Truth table construction and configuration analyses.** The variable calibration results were collapsed to obtain the set (i.e., the truth table) for each organization (i.e., each case) under *Coherence* and *Cognitive Participation* (4). The minimum frequency of occurrence of a solution (i.e., an antecedent variable) was set to 1 and the consistency of the solution was set to 0.8 based on the sample size included in the study (10,11). After the standard analysis, three scenarios for high-level of ORC configurations are obtained: Complex Solution, Intermediate Solution, and Parsimonious Solution (12). The intermediate and complex solutions in this study were in the same combination form, so they were merged and called equivalent solutions. The core conditions were identified through the nested relationship between the equivalent and parsimonious solution: Appearance in the equivalent and parsimonious solutions simultaneously were core conditions, and only in the equivalent solution were marginal conditions (12).

The solution consistency is Equationted as follows (3,10):

Equation 3:
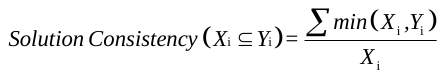

Where *X* is the solution, i.e., some configuration derived from this study, and *Y* is the outcome variable, i.e., high-level of ORC. *X*_i_ and *Y*_i_ are the membership of the solution and the outcome of the i case, i.e., the total number of membership for which the configuration achieves consistency divided by the total number of membership for that configuration. For cases that are inconsistent with the subset relationship, the part of the configuration where the membership is consistent with the result can be used to achieve this. min(*X*_i_, *Y*_i_) is the minimum of the two when *X* is compared with *Y*, i.e., when *X* takes a value greater than *Y*, the value of *Y* is brought into the calculation.

1. **Robustness test.** There were two approaches to the robustness test as an important step in QCA: based on Set-Theory and Statistical Theory (3). Given that the nature of QCA is based on the Set-Theory, the researcher suggested prioritizing the Set-Theory-based method (12). Based on the truth table and the literature, this study changed the consistency thresholds and added other variables related to the results for robustness tests to ensure that the results were reasonable and accurate (3,13).
2. **Interpret the result.** Based on condition variables composition characteristics, i.e., the presence and absence of core and marginal conditions, and combined with practical experiences, each configuration is specifically named to explain the internal mechanism, derive the path of high-level of ORC, and reveal the experiential model between the variables, which in turn provides practice-based guidance for the change implementation.

The study design and data analysis process in Appendix Figure 1.

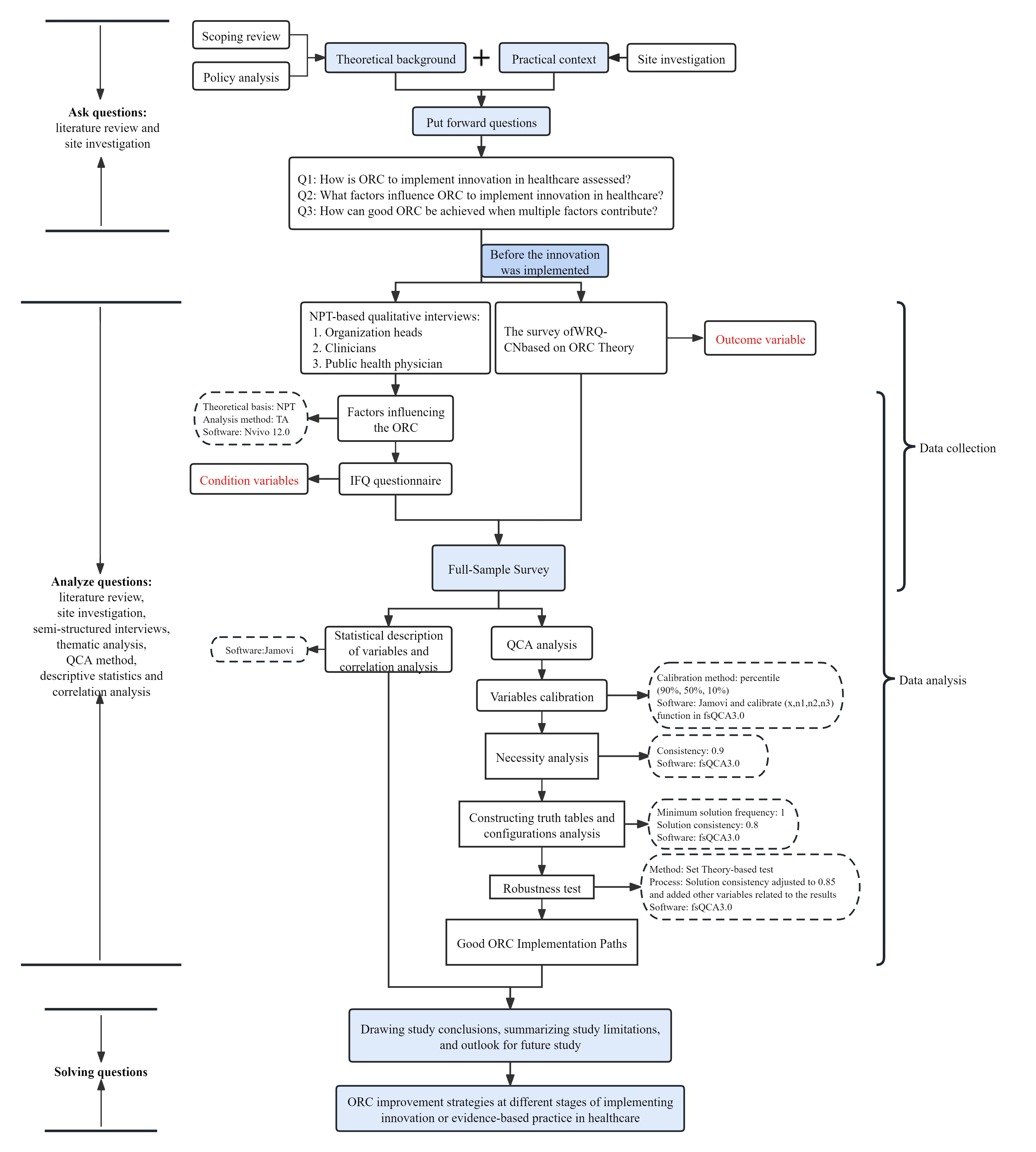

Appendix Figure 1 Study design and data analysis process

#### Appendix 6 Coding results for the condition variables

The interview results were coded using TA and categorized into eight second-level and 12 three-level indexes under *Coherence* and *Cognitive Participation*. The three-level indexes with the highest frequency of occurrence were Patient medication adherence and cognition], and the lowest frequency of occurrence was Professional knowledge] (Appendix Table 1).

Appendix Table 1 Thematic analysis coding results

| First-level indexes | Second-level indexes | Three-level indexes | Frequency statistics in text (times) |
| --- | --- | --- | --- |
| Coherence | Differentiation | Cost difference | 27 |
|  | Communal specification | Positive attitude | 40 |
|  | Individual specification | Specific tasks and responsibilities | 51 |
|  | Internalization | Value of innovation | 27 |
| Cognitive Participation | Initiation | Key participants | 44 |
|  | Enrolment | Preliminary preparation | 50 |
|  |  | Audit and feedback | 39 |
|  | Legitimation | Valid contribution for the innovation | 39 |
|  | Activation | Professional knowledge | 26 |
|  |  | Work pressure | 34 |
|  |  | Patient medication adherence and cognition | 62 |
|  |  | External supports | 30 |

#### Appendix 7 Descriptive statistics for condition variables and the ORC scores

| Variables | | | Frequency statistics for the choosing of the level of factor influence ^a^ | | | | | | Median | IQR ^b^ | | Max | Min |
| --- | --- | --- | --- | --- | --- | --- | --- | --- | --- | --- | --- | --- | --- |
|  |  |  | 0 | 1 | 2 | 3 | 4 | 5 |  | Q_L_ | Q_U_ |  |  |
| Outcome | | |  |  |  |  |  |  |  |  |  |  |  |
| ORC scores | | | - | - | - | - | - | - | 105.20 | 101.23 | 107.33 | 117.86 | 93.83 |
| Context | | | - | - | - | - | - | - | 34.10 | 33.28 | 36.25 | 41.14 | 28.50 |
| Change valence | | | - | - | - | - | - | - | 18.83 | 17.92 | 19.99 | 20.75 | 16.60 |
| Informational assessment | | | - | - | - | - | - | - | 18.71 | 18.25 | 19.10 | 21.00 | 16.80 |
| Change commitment | | | - | - | - | - | - | - | 17.67 | 16.66 | 18.75 | 21.00 | 15.60 |
| Change efficacy | | | - | - | - | - | - | - | 14.43 | 13.71 | 15.29 | 16.43 | 13.00 |
| Conditions | | |  |  |  |  |  |  |  |  |  |  |  |
| Coherence | Differentiation | Cost difference | 12 | 1 | 1 | 35 | 20 | 1 | 2.65 | 2.37 | 3.13 | 3.50 | 1.50 |
|  | Communal specification | Positive attitude | 7 | 4 | 3 | 28 | 25 | 3 | 3.10 | 2.36 | 3.34 | 3.83 | 2.14 |
|  | Individual specification | Specific tasks and responsibilities | 10 | 2 | 2 | 28 | 27 | 1 | 2.84 | 2.48 | 3.29 | 3.67 | 1.67 |
|  | Internalization | Value of innovation | 12 | 2 | 4 | 34 | 18 | 0 | 2.66 | 2.25 | 2.87 | 3.50 | 2.00 |
| Cognitive Participation | Initiation | Key participants | 6 | 3 | 3 | 26 | 27 | 5 | 3.07 | 2.71 | 3.50 | 3.83 | 2.17 |
|  | Enrolment | Preliminary preparation | 7 | 4 | 1 | 34 | 21 | 3 | 2.86 | 2.63 | 3.23 | 3.67 | 2.00 |
|  |  | Audit and feedback | 10 | 4 | 4 | 30 | 19 | 3 | 2.79 | 2.38 | 3.17 | 3.44 | 1.75 |
|  | Legitimation | Valid contribution to the innovation | 12 | 2 | 4 | 34 | 16 | 2 | 2.52 | 2.31 | 2.78 | 3.67 | 2.00 |
|  | Activation | Professional knowledge | 7 | 4 | 1 | 23 | 29 | 6 | 3.17 | 2.97 | 3.54 | 4.00 | 2.29 |
|  |  | Work pressure | 11 | 4 | 1 | 30 | 20 | 4 | 2.75 | 2.55 | 2.96 | 4.00 | 1.75 |
|  |  | Patient medication adherence and cognition | 6 | 3 | 2 | 23 | 27 | 9 | 3.29 | 3.00 | 3.53 | 3.83 | 2.29 |
|  |  | External supports | 5 | 5 | 0 | 24 | 27 | 9 | 3.31 | 3.06 | 3.53 | 4.00 | 2.40 |

*Note:* a: 0 = no influence, 1 = very low influence, 2 = relatively low influence, 3 = fair influence, 4 = relatively high influence, 5 = very high influence

b: Q_L_: lower quartile, Q_U_: upper quartile

#### Appendix 8 Correlation analysis

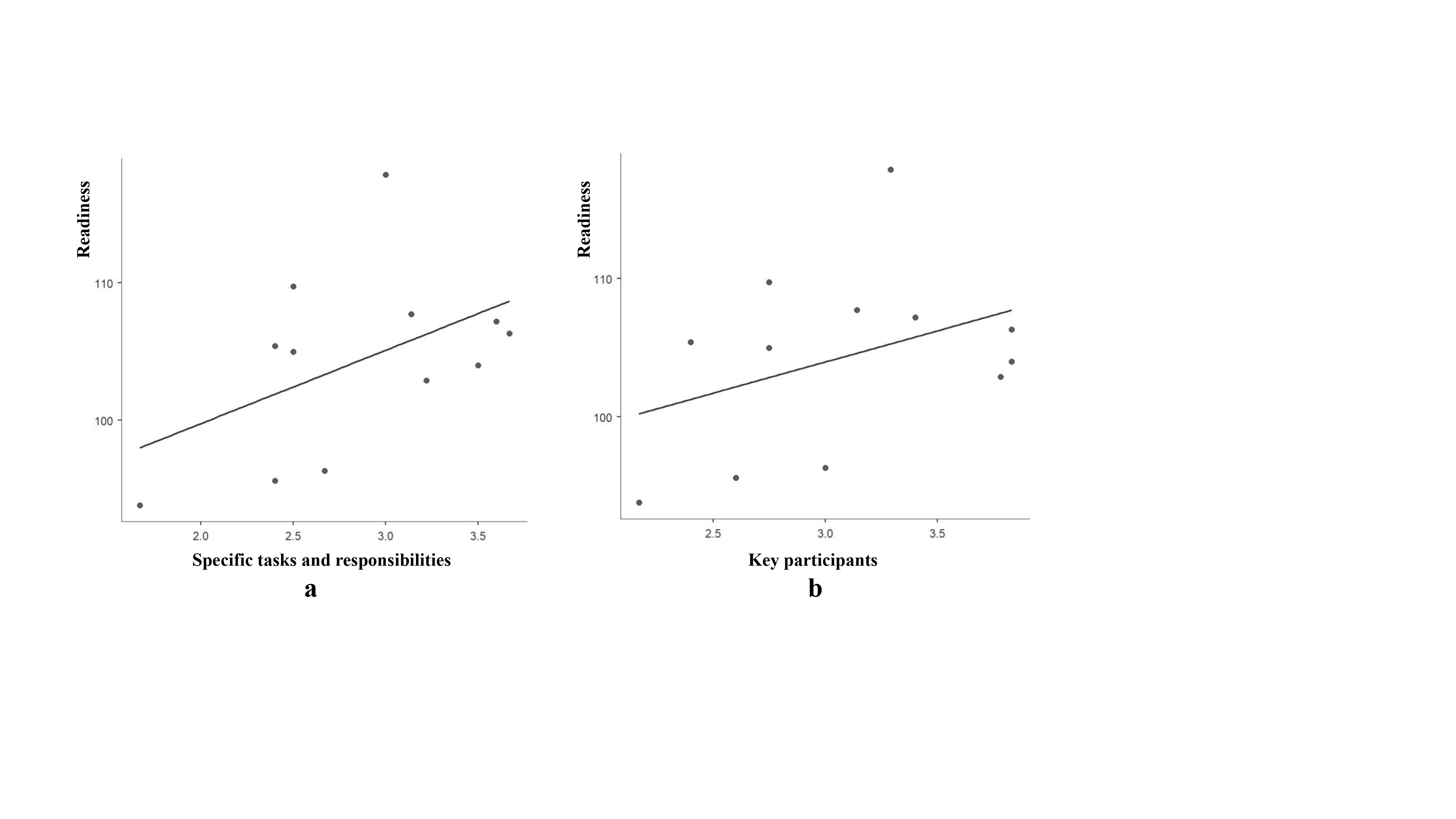

#### Appendix 9 Variables calibration and Necessity analysis

The percentile method calibrated the data uniformly (1), facilitating the overall consistency and coverage of the analysis. The calibration results showed that the best consistency and coverage were achieved when the calibration points were 90%, 50%, and 10%. The ORC score is ≥ 115.43, it was full membership with the high ORC set; when 105.20 ≤ ORC score < 115.43, it was strong membership with the high-level ORC set; When 94.36 ≤ ORC score < 105.20, it was weakly membership with the high ORC set; when ORC score < 94.36, it was full nonmembership with the high ORC set. The remaining variables are membership and so on. The specific calibrations are shown in Appendix Table 1.

Based on Equation 1 and variable calibration results, this study calculated the membership of each organization on each variable as detailed in Appendix Table 2.

Appendix Table 1 Variables calibration results

| Variables | | | Full membership | Crossover points | Full nonmembership |
| --- | --- | --- | --- | --- | --- |
| Outcome | ORC scores | | 115.43 | 105.20 | 94.36 |
| conditions | | |  |  |  |
| Coherence | Differentiation | Cost difference | 3.44 | 2.65 | 1.65 |
|  | Communal specification | Positive attitude | 3.82 | 3.10 | 2.15 |
|  | Individual specification | Specific tasks and responsibilities | 3.65 | 2.84 | 1.89 |
|  | Internalization | Value of innovation | 3.35 | 2.66 | 2.00 |
| Cognitive Participation | Initiation | Key participants | 3.83 | 3.07 | 2.24 |
|  | Enrolment | Preliminary preparation | 3.60 | 2.86 | 2.12 |
|  |  | Audit and feedback | 3.37 | 2.79 | 1.77 |
|  | Legitimation | Valid contribution for the innovation | 3.53 | 2.52 | 2.04 |
|  | Activation | Professional knowledge | 3.94 | 3.17 | 2.38 |
|  |  | Work pressure | 3.88 | 2.75 | 1.86 |
|  |  | Patient medication adherence and cognition | 3.82 | 3.29 | 3.32 |
|  |  | External supports | 3.93 | 3.31 | 2.43 |

Appendix Table 2 Organization membership for each variable

| Variables | | | A | B | C | D | E | F | G | H | I | J | K | L |
| --- | --- | --- | --- | --- | --- | --- | --- | --- | --- | --- | --- | --- | --- | --- |
| Outcome | ORC scores | | 0.98 | 0.51 | 0.07 | 0.08 | 0.42 | 0.58 | 0.68 | 0.64 | 0.49 | 0.79 | 0.04 | 0.35 |
| Conditions | | | | | | | | | | | | | | |
| Coherence | Differentiation | Cost difference | 0.34 | 0.21 | 0.64 | 0.86 | 0.88 | 0.96 | 0.92 | 0.12 | 0.03 | 0.39 | 0.39 | 0.85 |
|  | Communal specification | Positive attitude | 0.69 | 0.10 | 0.60 | 0.05 | 0.84 | 0.95 | 0.05 | 0.60 | 0.06 | 0.42 | 0.13 | 0.94 |
|  | Individual specification | Specific tasks and responsibilities | 0.64 | 0.20 | 0.20 | 0.37 | 0.92 | 0.96 | 0.75 | 0.94 | 0.25 | 0.25 | 0.02 | 0.80 |
|  | Internalization | Value of innovation | 0.55 | 0.05 | 0.43 | 0.18 | 0.68 | 0.97 | 0.55 | 0.05 | 0.81 | 0.05 | 0.18 | 0.81 |
| Cognitive Participation | Initiation | Key participants | 0.70 | 0.08 | 0.15 | 0.44 | 0.95 | 0.95 | 0.57 | 0.79 | 0.24 | 0.24 | 0.04 | 0.94 |
|  | Enrolment | Preliminary preparation | 0.50 | 0.11 | 0.02 | 0.78 | 0.87 | 0.96 | 0.50 | 0.80 | 0.16 | 0.38 | 0.29 | 0.91 |
|  |  | Audit and feedback | 0.59 | 0.24 | 0.05 | 0.21 | 0.88 | 0.88 | 0.44 | 0.89 | 0.05 | 0.47 | 0.55 | 0.97 |
|  | Legitimation | Valid contribution for the innovation | 0.08 | 0.33 | 0.56 | 0.24 | 0.61 | 0.97 | 0.37 | 0.88 | 0.04 | 0.16 | 0.61 | 0.85 |
|  | Activation | Professional knowledge | 0.23 | 0.10 | 0.34 | 0.34 | 0.50 | 0.88 | 0.03 | 0.92 | 0.96 | 0.78 | 0.50 | 0.65 |
|  |  | Work pressure | 0.57 | 0.13 | 0.97 | 0.30 | 0.75 | 0.43 | 0.35 | 0.91 | 0.15 | 0.03 | 0.55 | 0.59 |
|  |  | Patient medication adherence and cognition | 0.29 | 0.06 | 0.95 | 0.77 | 0.56 | 0.95 | 0.04 | 0.85 | 0.77 | 0.47 | 0.29 | 0.29 |
|  |  | External supports | 0.36 | 0.04 | 0.80 | 0.38 | 0.52 | 0.97 | 0.48 | 0.61 | 0.71 | 0.06 | 0.16 | 0.91 |

*Note:* a: The 12 institutions included in the study are denoted by A to L.

The results showed that when the outcome variable is high ORC, under the Coherence, Specific tasks and responsibilities have the highest consistency result of 0.75; under the Cognitive Participation, Key participants and Audit and feedback have the highest consistency results of 0.71 and 0.70, respectively. However, the consistency of all condition variables was less than 0.9. None of the condition variables constituted a necessary condition to produce a high ORC (Appendix Table 3).

Appendix Table 3 Results of the necessity analysis

| Condition variables | | | High-level readiness | |
| --- | --- | --- | --- | --- |
| First-level indexes | Second-level indexes | Three-level indexes | Consistency | Coverage |
| Coherence | Differentiation | Cost difference | 0.59 | 0.50 |
|  | ~Differentiation | ~Cost difference | 0.62 | 0.65 |
|  | Communal specification | Positive attitude | 0.61 | 0.63 |
|  | ~Communal specification | ~Positive attitude | 0.61 | 0.52 |
|  | Individual specification | Specific tasks and responsibilities | 0.75 | 0.66 |
|  | ~Individual specification | ~Specific tasks and responsibilities | 0.52 | 0.51 |
|  | Internalization | Value of innovation | 0.59 | 0.62 |
|  | ~Internalization | ~ value of innovation | 0.66 | 0.56 |
| Cognitive Participation | Initiation | Key participants | 0.71 | 0.66 |
|  | ~Initiation | ~Key participants | 0.54 | 0.52 |
|  | Enrolment | Preliminary preparation | 0.67 | 0.60 |
|  | ~Enrolment | ~Preliminary preparation | 0.58 | 0.57 |
|  |  | Audit and feedback | 0.70 | 0.64 |
|  |  | ~Audit and feedback | 0.55 | 0.53 |
|  | Legitimation | Valid contribution for the innovation | 0.56 | 0.55 |
|  | ~Legitimation | ~Valid contribution for the innovation | 0.75 | 0.67 |
|  | Activation | Professional knowledge | 0.68 | 0.61 |
|  | ~Activation | ~Professional knowledge | 0.60 | 0.59 |
|  |  | Work pressure | 0.58 | 0.57 |
|  |  | ~Work pressure | 0.76 | 0.68 |
|  |  | Patient medication adherence and cognition | 0.62 | 0.55 |
|  |  | ~Patient medication adherence and cognition | 0.67 | 0.67 |
|  |  | External supports | 0.64 | 0.60 |
|  |  | ~External supports | 0.69 | 0.65 |

#### Appendix 10 Robustness test results with different conditions

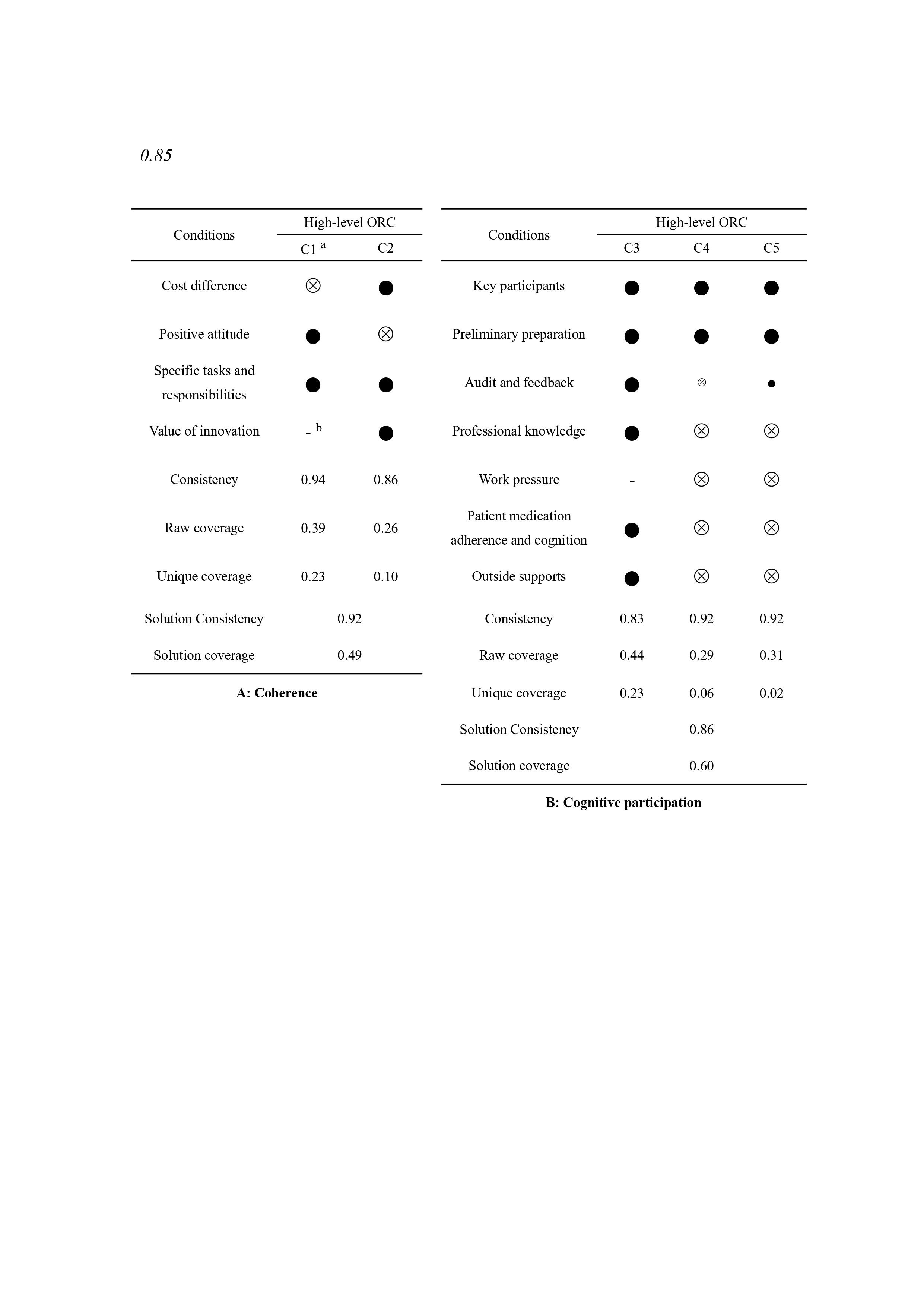

A: Configuration analysis based on 0.85 consistency value

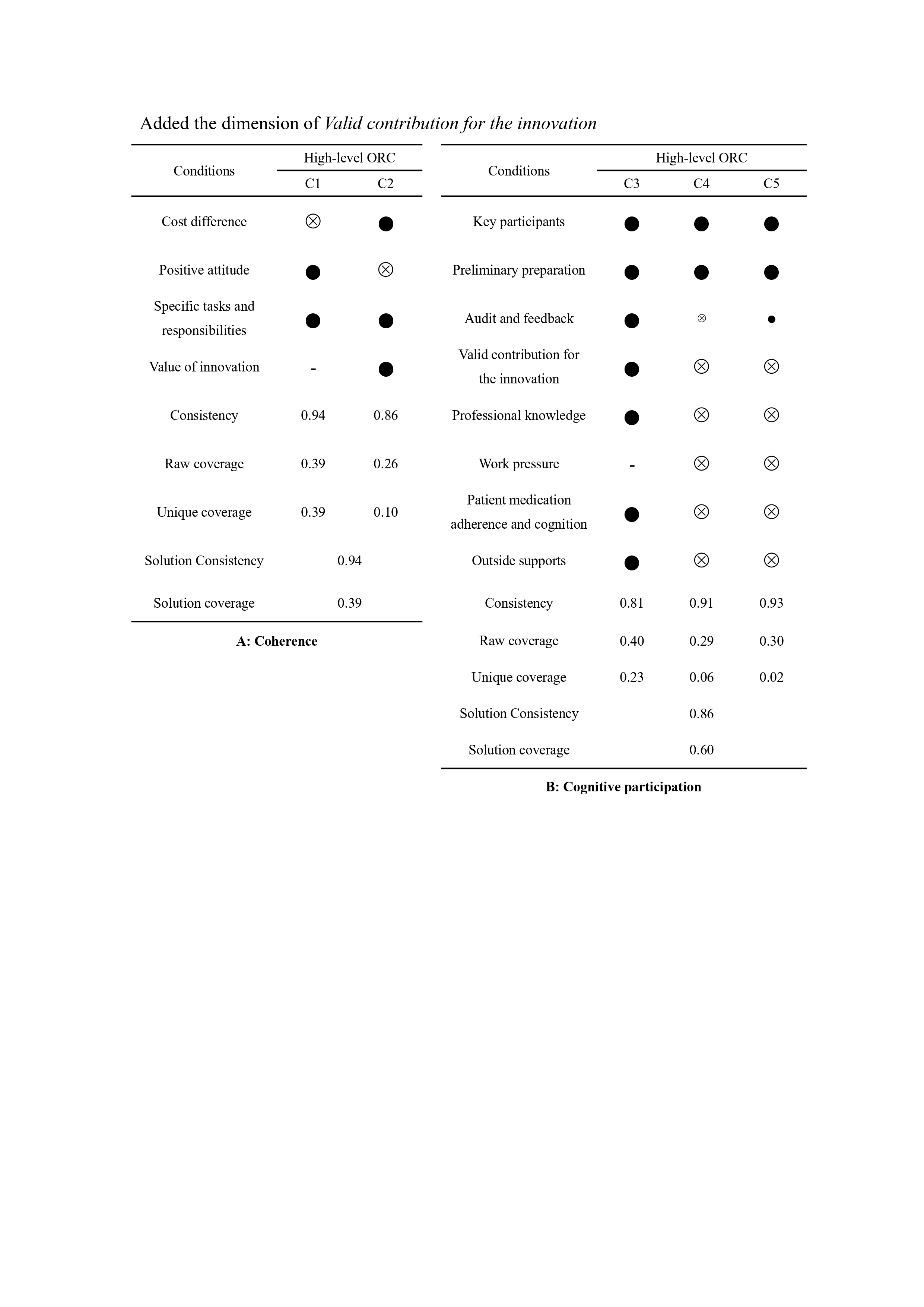

**B: Configuration analysis based on adding the *Valid contribution for the innovation* variable**

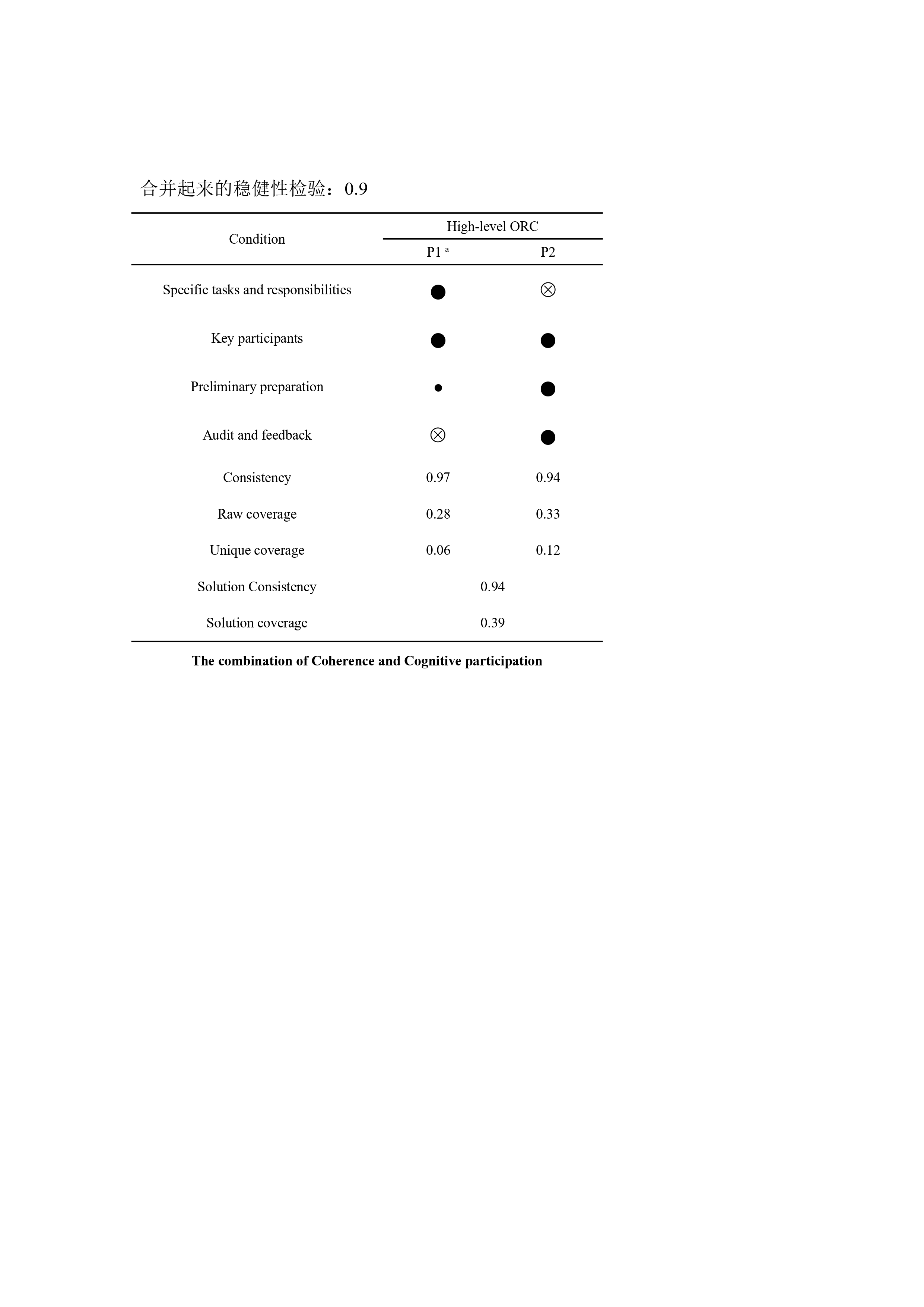

C: The robustness of pathway analysis

*Note:*

In this study, Ragin and Fiss's variable labeling methods (1) were used:

● /⊗= Core conditions present/absent; ●/⊗ = Marginal conditions present/absent；

a: C/P: The abbreviation of configuration/pathway.

b: ["-"] indicates that the variable does not work in the configuration
